# Imaging Drug Resistance in Juvenile Myoclonic Epilepsy with MRI-derived Cortical Markers

**DOI:** 10.1101/2023.12.03.23298783

**Authors:** Bernardo Crespo Pimentel, Giorgi Kuchukhidze, Fenglai Xiao, Lorenzo Caciagli, Julia Höfler, Lucas Rainer, Martin Kronbichler, Christian Vollmar, John S Duncan, Eugen Trinka, Matthias Koepp, Britta Wandschneider

## Abstract

Juvenile myoclonic epilepsy (JME) is the most common idiopathic generalized epilepsy (IGE) syndrome and can present a significant clinical challenge due to lack of seizure control and concomitant neuropsychological impairment. Its underlying mechanisms remain poorly understood. MRI studies have described structural changes in individuals with IGE syndromes but have so far not identified a distinct structural profile associated with drug resistant IGE. In this cross-sectional case-control neuroimaging analysis we included 79 participants with JME recruited at two European epilepsy centres, 43 of whom were drug resistant, as defined by recurrent seizures in the last 12 months despite adequate trials of at least two antiseizure medications (ASM), a drug sensitive group (*n*=37), and 78 healthy controls. Participants underwent T1-weighted MRI and a comprehensive neuropsychological test battery including measures within two different domains: verbal memory and executive function. We performed vertex-wise measurements of cortical thickness and neurodevelopmental cortical measures, i.e. white matter surface area and local gyrification index (LGI), through a surface-based framework with subsequent correction of scanner-related effects. Group comparisons (family-wise error (FWE) corrected at *p* <0.05, family-wise error corrected) showed an increase of surface area in prefrontal, cingulate and temporal regions with left-sided predominance accompanied by increased LGI in the left temporal lobe in drug-resistant compared to drug-responsive individuals with JME. Performance on executive function in participants with drug resistant JME was reduced compared to drug sensitive JME (*p*_Bonferroni_ <0.01) and controls (*p*_Bonferroni_ <0.001). Impaired executive function correlated with increased surface area in medial prefrontal and temporal cortical regions (*r* < -0.75, *p* <0.01) after adjusting for disease duration and total ASM load. In conclusion, we identified a cognitive-developmental profile in drug resistant JME characterized by left-weighted changes in surface area and cortical folding complexity, the extent of which correlates with the degree of executive dysfunction. Our results shed further light onto the pathophysiological mechanisms of JME and suggest a neurodevelopmental/network basis for both drug refractoriness and cognitive impairment.

## Introduction

Juvenile myoclonic epilepsy (JME) is the most common idiopathic generalized epilepsy (IGE) syndrome, affecting 5-10% of all people with epilepsy^1^. Good treatment response was long regarded as a core characteristic of IGE syndromes. This feature was dropped from the ILAE 2022 classification of epilepsy syndromes^2^ in an attempt to include drug resistant individuals under the same umbrella.^3^ Similar to focal epilepsies, up to 30% of people JME are refractory to anti-seizure medications (ASM).^4–8^ Individuals with drug resistant IGE syndromes also tend to present with seizures at a younger age and have a higher incidence of psychiatric comorbidity and cognitive impairment, mainly affecting frontal lobe functions such as working memory and executive function.^6,9–14^ By definition, clinical imaging is unremarkable in IGE. Multiple large-scale MRI and genetic studies have not yet supported the concept of a distinct structural phenotype associated with drug resistance.^15–17^ Such studies have, however, mostly focused on individuals with IGE syndromes in which seizures persist into adulthood and thus represent cohorts who are unlikely to achieve disease remission.

Quantitative MRI measures of cortical thickness and other brain surface metrics deliver robust and reproducible markers of neurodegeneration and neurodevelopment.^18–20^ Neuroimaging studies in JME mainly identified structural anomalies in thalamic, premotor, and medial prefrontal areas^21–29^, but also more recently including the mesiotemporal lobe.^30–32^ It remains unclear whether such abnormalities represent disease developmental signatures or are rather a consequences of the disease. The concept of endophenotype (i.e., disease trait that is more prevalent in patients and first-degree relatives than the general population) has been used in imaging studies in to disentangle characteristics attributable to the genetic underpinnings of disease from the effects of disease activity or ASM. Notably, in a recent endophenotype study we identified changes in surface area and sulcal-gyral complexity as neurodevelopmental markers in JME.^19^ As cortical surface characteristics are believed to be highly heritable^33^, we hypothesize that alterations of cortical architecture may reflect a continuum determined by different degrees of genetic vulnerability and manifesting in distinct clinical phenotypes, possibly explaining intractability in JME.

Here, we aimed to identify cortical structural correlates of drug resistance in a large multicentric JME cohort. We used structural MRI to provide quantitative measures of cortical organization and folding that reflect brain development. Neuropsychological tests were implemented to address verbal memory and executive function and thus characterize the cognitive phenotype of drug-resistant JME. We then assessed the relationship between domain-specific cognitive performance and changes in cortical surface features to identify developmental alterations that underpin both disease severity and cognitive impairment.

## Materials and methods

### Participants

We included a total of 79 individuals with JME recruited from the epilepsy outpatient clinics at the Department of Neurology, Christian Doppler University Hospital in Salzburg, Austria (CDK) (51 patients, 59 controls) and University College London Hospitals, London, UK (UCL) (28 patients, 19 controls) between 2007 and 2019. People with JME were divided into a drug resistant (*n*=42) and a drug responsive group (*n*=37). Drug resistance was defined as ongoing seizures in the 12 months prior to MRI acquisition despite at least two appropriate and well-tolerated ASM trials.^34^ Seventy-eight healthy controls comparable for age and sex were included for comparisons. All participants had an otherwise unremarkable neurological history. MRI scans were reported as normal by a neuroradiologist. Results on cortical morphology were previously reported for part of the JME cohort and healthy controls.^19^

Apart from clinical and demographical metrics, we quantified the total ASM load by calculating the ratio of the actual daily dose of a specific ASM by its defined daily dose (DDD), provided by the Collaborating Centre for Drug Statistics Methodology of the World Health Organization, as previously reported^35^. The cumulative ASM load for each individual is the sum of this ratio for all ASMs of the individual drug regimen.

All study procedures were carried out in accordance with the Declaration of Helsinki. The study was approved by the corresponding ethics committee of the UCL Queen Square Institute of Neurology and University College London Hospitals as well as the region of Salzburg. All participants provided written informed consent.

### Data acquisition and preprocessing

Structural MRI data at the CDK were obtained with a 3T Magnetom Prisma-fit 3-T MRI scanner using a 3D multiecho MPRAGE sequence (repetition time 2.4ms, echo time 2.2ms, inversion time 1060ms, flip angle = 8°, matrix = 320×300, voxel size = 0.8×0.8×0.8mm^3^). Participants from UCL were scanned using a GE MR750 3T MRI scanner using a 3D fast spoiled gradient echo sequence (repetition time 7.2ms, echo time 2.8ms, inversion time 450ms, flip angle = 20°, matrix = 256×256, voxel size = 1.1×1.1×1.1mm^3^). Both MRI datasets were processed with the standardized Freesurfer processing pipeline.^36^ The processing stream consisted of automated transformation to Talairach space, skull stripping, intensity normalization and segmentation of white/grey matter tissue, resulting in extraction of surface meshes composed of approximately 150,00 vertices in each hemisphere. Surface extractions were visually assessed and manually corrected. Individual surfaces were then registered to an average template surface.

### Computation of surface parameters

*Cortical thickness* was calculated as the distance between corresponding vertices in the white matter and pial surfaces. In line with previous work, *surface area* was calculated based on the average area of 6 triangles surrounding the index vertex on the white matter surface, thus reflecting the vertex-wise degree of cortical expansion or compression.^37^ *Local gyrification Index* (LGI) was computed at each vertex using an inbuilt Freesurfer function which quantifies the gyrification index in a three-dimensional framework.^38^ LGI represents the amount of cortex buried within the sulcal folds compared with the amount of visible cortex across the whole cortical surface in each vertex. Cortex with extensive folding has a large gyrification index, whereas a cortex with limited folding has a small gyrification index.^39–42^ Cortical thickness and surface area were smoothed with a 20 mm surface-based kernel, whereas a kernel of 5 mm was applied to LGI due to prior inherent smoothing in metric calculation. All metrics were resampled to a template surface. Prior to downstream statistical analysis, cortical thickness, surface area and LGI measurements was corrected for scanner-related batch effects using the Combat Tool for Harmonization of Multi-Site Imaging Data in R.^43^ Combat is a validated harmonisation tool for multi-scanner cross-sectional comparison of cortical thickness^44^ and surface area^43^ measurements, among others.

### Neuropsychological evaluation

All participants underwent comprehensive neuropsychological assessment in their primary language on the same or the day after scanning. Evaluations included standardized measures of executive function and verbal learning as well as self-reported questionnaires assessing mood (depression and/or anxiety). Because of discrepancies in the tests employed in both centres, patients’ raw scores were standardized (*z*-scores) in relation to the control group of the respective centre. Supplementary Table 1 provides a list of used cognitive tests and self-assessment questionnaires.

### Statistical analysis

Demographic and clinical data were analyzed with R studio (version 2023.03.0+386). Kruskal-Wallis test was used to compare continuous clinical characteristics between subgroups. Pearson’s Chi-Square was used for categorical data. *Post-hoc* tests were performed using Bonferroni-corrected pairwise comparisons. All statistic tests were performed two-tailed.

#### Surface-based exploratory analysis

Statistical analysis was carried out using BrainStat for Matlab.^45^ We first explored the effect of age (in the whole cohort) as well as disease duration (in the patient group) on the different surface features in order to account for their contribution to the group-comparison models (see eFig. 1) with vertex-wise two-tailed student t-tests. Cortical thickness, surface area and LGI were then compared between the patient and control group (to look for disease related effects), as well as between the drug resistant and drug sensitive group (to look for effects related to disease severity) with vertex-wise two-tailed Student t-tests. Here, a binary indicator of group (e.g., 0 = healthy controls, 1 = person with drug resistant epilepsy) was the predictor of interest, whereas cortical thickness, surface area or LGI of a specific vertex was the outcome of interest. We calculated vertex-wise effect size estimates using Cohen’s *d*. After verification of age and disease duration effect on the different cortical metrics, cortical thickness and LGI were controlled for age and sex, whereas surface area was adjusted for sex and total white matter volume. In order to consider the effects of brain maturation on surface area variation, we performed an additional analysis in which surface area was also corrected for age. We applied multiple comparison correction using random field theory^46^ (RFT) at a family-wise-error (FWE) of 0.05.

**Figure 1.**
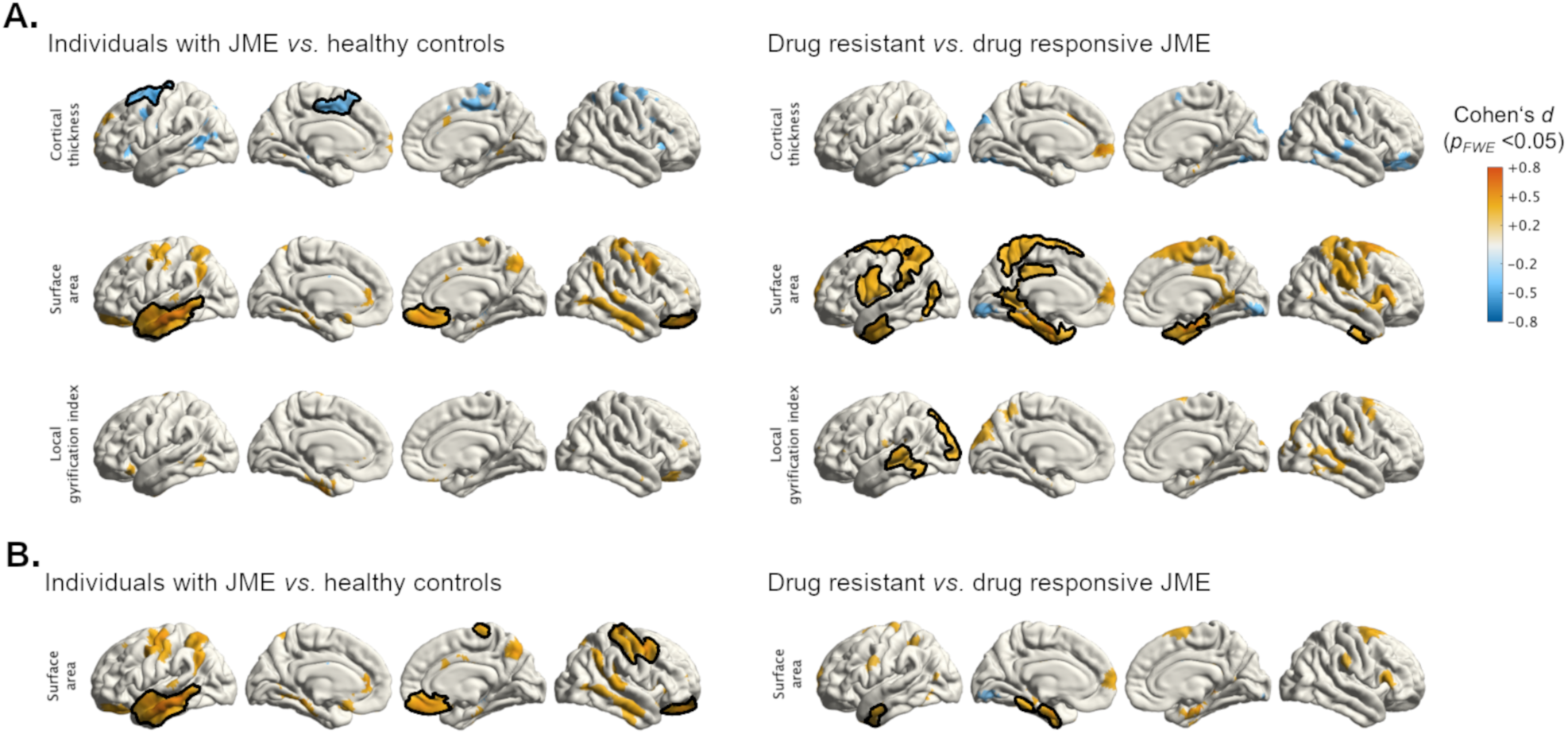
Effects on cortical surface related to syndrome and disease severity. (A) Mass univariate analysis showing group comparisons of cortical thickness, surface area and local gyrification index (LGI) between individuals with JME and healthy controls (syndrome-related effects) and between drug resistant and drug responsive individuals with JME (effects related to disease severity). Cortical thickness and LGI were adjusted for age and sex, whereas surface area was only corrected for sex and total white matter volume (see Methods). (B) shows group comparisons of surface area after adjusting for the effect of age as a surrogate marker of brain maturation. Clusters are color-coded according to the corresponding effect size estimates as reported by Cohen’s *d* (see color bar). Clusters that survived multiple comparisons correction using random field theory at *p*_FWE_ <0.05 are outlined in black.

#### Neuropsychological data

We used multivariate analysis of covariance (MANCOVA) to compare neuropsychological data between groups and included age and sex as covariates while using Wilk’s lambda (λ) as multivariate test statistic. For all neuropsychological tests, we compared drug resistant patients, drug sensitive patients and healthy controls by performing univariate analysis of covariance (ANCOVA) on the z-standardized test scores while adjusting for age as a continuous variable as well as and sex and centre (CDK or UCL) as dichotomous variables. ANCOVA *p*-values were corrected for multiple comparisons using false discovery rate (FDR). Pairwise t-tests on residualized metrics (hence age- and sex-adjusted) were Bonferroni-corrected for multiple comparisons. Missing data were addressed via pairwise deletion in all analysis. Eta-square (η²) was used as a measure of effect-size. We repeated the ANCOVA additionally adjusting for hospital anxiety and depression (HADS-A & HADS-D) scores.

To reduce data dimensionality, two principal component analyses (PCA) were run and composite cognitive variables were obtained for: (i) executive function (TMT B-A, digit span, lexical fluency, semantical fluency) and (ii) verbal memory (list learning and list recall). We included language fluency within the scope of executive function, as previously described.^47^ The first principal component of each PCA was retained for further correlation analysis after verification that it explained a sizeable amount of the variance (>40%) and had an eigenvalue >1.

To explore the influence of disease chronicity on cognition, we conducted correlation analysis between the composite cognitive scores of executive function and verbal memory with disease duration across all patients. We also performed correlation analysis of cognitive domain scores with age at onset. Because age at onset and disease duration were collinear (*r* = -0.24, *p* = 0.05), we opted for partial correlations of composite cognitive scores with age at onset covaried for disease duration and vice-versa. Due to the influence of ASM on cognition, we used the total ASM load score as a covariate in all partial correlations.

#### Correlation between cognitive domains and brain anatomy

To explore the relationship between brain surface architecture and executive function/verbal memory, we performed a vertex-wise partial *Pearson* correlation analysis between each brain surface feature and the composite cognitive score for executive function verbal memory for each participant subgroup (controls, drug resistant and drug sensitive participants). We adjusted each correlation analysis for disease duration and total ASM score in both the drug-resistant and drug sensitive group, and for age in the control group. Similar to our group comparison analysis, we performed multiple comparison correction with RFT^46^ at *p*_FWE_ <0.05.

## Results

Healthy controls and people with JME were comparable in age (median [IQR (Interquartile Range)]: 26 years [22-32] vs. 28 years [24-35], *p* = 0.06), sex [females: 44(56%) vs. 43(54%), *p* = 0.92] and center affiliation [from CDK Salzburg: 59/78 (76%) controls, 51/79 (65%) people with JME, *p* = 0.13]. Although individuals with drug resistant disease, drug responsive disease and healthy controls were not comparable for age (Kruskal-Wallis *p* <0.01), *post-hoc* testing revealed only a significant difference between controls and drug responsive participants (median [IQR]: 26 years [22, 32] vs. 32 years [25, 38], respectively; *p*_Bonferroni_ <0.05). Participants with drug resistant JME presented with a shorter disease duration (median [IQR]: 12 years [8, 19] vs. 16 years [13, 24], Kruskal-Wallis *p* <0.05) and a higher total number of ASM trials in life (median [IQR]: 2.00 [2.00, 2.00] vs. 2.00 [1.00, 2.00], Kruskal-Wallis *p* <0.001) than drug sensitive participants. Both disease subgroups expressed higher rates of anxiety (median [IQR]: healthy controls = 3.0 [1.8, 5.0] vs. drug resistant = 5.0 [3.8, 8.5] vs. drug sensitive = 5.0 [2.0, 7.0]; Kruskal-Wallis *p* <0.001) and depressive symptoms (median [IQR]: healthy controls = 2.0 [1.0, 3.0] vs. drug resistant = 4.0 [2.0, 6.0] vs. drug responsive = 2.0 [1.0, 5.0]; Kruskal-Wallis *p* <0.001). All groups were comparable for sex, ASM total drug load and prevalence of bilateral tonic-clonic seizures. Demographic characteristics of study participants are provided in Table 1. Comparisons between controls and all people with JME are provided in Supplementary Table 1.

**Table 1.** Demographics, clinical characteristics and self-assessment questionnaires.

|  | Controls<br>(n = 78) | Drug resistant<br>(n = 42) | Drug sensitive<br>(n = 37) | Test<br>statistic | p value | Post-hoc p values<br>(Bonferroni corrected) |
| --- | --- | --- | --- | --- | --- | --- |
| **Demographics & clinical characteristics** |  |  |  |  |  |  |
| Age, median [IQR], y |  |  |  |  |  |  |
| At MRI scan | 26 [22, 32] | 26 [22, 32] | 32 [25, 39] | 10.97<sup>a</sup> | **<0.01** | Controls/drug resistant: 0.90<br>Controls/drug sensitive: **<0.05**<br>Drug resistant/drug sensitive: 0.09 |
| At seizure onset |  | 14.0 [12.0, 16.8] | 15.0 [13.0, 16.0] | 0.3<sup>a</sup> | 0.549 | - |
| Female, No. (%) | 44 (56) | 21 (50) | 22 (59) | 0.77<sup>b</sup> | 0.679 | - |
| Duration of epilepsy, median [IQR], y |  | 12 [8, 19] | 16 [13, 24] | 6.07 | **<0.05** | - |
| Days since last seizure, median [IQR], d |  | 95 [11, 284] | 1500 [660, 2445] | 43.56 | **<0.001** | - |
| Total Number of ASM trials, median [IQR] |  | 2.00 [2.00, 2.00] | 2.00 [1.00, 2.00] | 11.35 | **<0.001** | - |
| Total ASM load score, median [IQR] |  | 1.20 [0.67, 1.97] | 1.00 [0.40, 1.33] | 0.63 | 0.086 | - |
| Bilateral tonic-clonic seizures, No. (%) |  | 39 (93) | 28 (76) | 3.27 | 0.057 | - |
| Currently on zonisamide/topiramate, No. (%) |  | 7 (17) | 4 (11) | 0.18 | 0.528 | - |
| **Self-assessment questionnaires** |  |  |  |  |  |  |
| HADS-A, median [IQR], score | 3.0 [1.8, 5.0] | 5.0 [3.0, 8.5] | 5.0 [2.0, 7.0] | 16.07<sup>a</sup> | **<0.001** | Controls/drug resistant: **<0.001**<br>Controls/drug sensitive: **0.01**<br>Drug resistant/drug sensitive: 0.90 |
| HADS-D, median [IQR], score | 2.0 [1.0, 3.0] | 4.0 [2.0, 6.0] | 2.0 [1.0, 5.0] | 17.61<sup>a</sup> | **<0.001** | Controls/drug resistant: **<0.001**<br>Controls/drug sensitive: **<0.05**<br>Drug resistant/drug sensitive: 0.90 |

Abbreviations: ASM, anti-seizure medication; HADS-A/D, hospital anxiety and depression scale – anxiety/depression IQR, interquartile range; MRI, magnetic resonance imaging; PMU, Paracelsus Medical University; <sup>a</sup>Kruskal-Wallis test, <sup>b</sup>Pearson's $\chi^2$
Note: bold indicates statistical significance.

### Analysis of cortical markers

#### Effect of age and disease duration on cortical markers

The association of age and disease duration on cortical markers is displayed in Supplemental Fig. 1. Both age and disease duration showed a significant negative association on cortical thickness and LGI in widespread cortical areas, that is, thinner cortex and lower LGI with increasing age. No statistically significant effect was seen for white matter surface area.

#### Individuals with JME vs. healthy controls

Compared to healthy controls, the JME group showed cortical thinning in the medial and superior lateral aspect of the left superior frontal gyrus. This group also presented with significantly increased surface area in the anterolateral aspect of the left temporal lobe as well as in the right orbitofrontal cortex (Fig. 1A). When adding age as a covariate to the surface area model, an additional cluster comprising the right pre- and postcentral gyrus gained significance (Fig. 1B). No changes in LGI were observed between patients and healthy controls.

#### Individuals with drug resistant vs. drug sensitive JME

Except for a discrete cluster of cortical thinning in the left inferior temporal gyrus in drug resistant JME, we observed no significant abnormalities in cortical thickness between the groups. Individuals with drug resistant JME displayed increased white matter surface area in multiple areas of the medial and lateral aspects of the left prefrontal and parietal cortices as well as the medial temporal lobes (left more than right) and inferior aspect of the temporal pole bilaterally (Fig. 1A). When correcting for age as a marker of brain maturation, only discrete areas in the left temporal lobe survived family-wise correction (Fig. 1B). LGI was increased in the left lateral temporal lobe and occipital cortex in drug resistant individuals (Fig. 1A). Repeat patient subgroup analysis additionally covarying for total ASM load did not alter the results.

### Cognitive performance across groups

#### Neuropsychological test *z*-scores

Complete neuropsychological assessment was available for a total of 69 participants with JME (36 of whom had drug resistant and 33 drug sensitive disease) and 59 healthy controls. MANCOVA covarying for age, sex and centre yielded a significant effect of group for cognitive performance (Wilk’s lambda = 0.65, *F* = 3.90, *p* <0.001). Subsequent ANCOVAs adjusted for sex, age and centre demonstrated significant group differences in TMT B-A (*p*_FDR_ < 0.01), TMT A (*p*_FDR_ < 0.001), digit span backwards (*p*_FDR_ < 0.01), verbal learning (*p*_FDR_ *<* 0.01), letter fluency (*p*_FDR_ = 0.001), and word fluency (*p*_FDR_ = 0.001). Repeat analysis additionally correcting for either anxiety or depression self-assessment scores did not alter the significance of results. Post-hoc tests showed worse performance of drug resistant patients against controls in all cognitive tests except for verbal recall (for letter fluency and trail-making test (TMT) A, *p*_Bonferroni_ < 0.001; for digit span backwards, word fluency and verbal learning *p*_Bonferroni_ < 0.01; for TMT B-A, *p*_Bonferroni_ = 0.05). This group also performed worse than drug sensitive individuals in the TMT A test (*p*_Bonferroni_ *=* 0.001), digit span backwards (*p*_Bonferroni_ < 0.05), letter fluency (*p*_Bonferroni_ < 0.05) and word fluency (*p*_Bonferroni_ < 0.01). Detailed statistics regarding neuropsychological test outcomes are provided in Table 2.

**Table 2.** Performance in neuropsychological tests across groups (*z*-scores)

|  | Drug resistant<br>(*n* = 36) | Drug responsive<br>(*n* = 33) | F Statistic | *p*<sub>FDR</sub> value | Post hoc *p* values*<br>(Bonferroni corrected) |
| --- | --- | --- | --- | --- | --- |
| **Executive function** |  |  |  |  |  |
| TMT B-A | 1.00 (0.32) | 0.66 (0.25) | 6.77 | **<0.01** | Controls/drug resistant: **<0.05**<br>Controls/drug responsive: > 0.90<br>Drug resistant/drug responsive: 0.20 |
| Digit span backwards | -0.46 (0.15) | 0.30 (0.15) | 6.39 | **<0.01** | Controls/drug resistant: **<0.01**<br>Controls/drug responsive: 0.90<br>Drug resistant/drug responsive: **<0.05** |
| Letter fluency | -0.91 (0.19) | -0.30 (0.18) | 8.88 | **0.001** | Controls/drug resistant: **<0.001**<br>Controls/drug responsive: 0.60<br>Drug resistant/drug responsive: **<0.05** |
| Word fluency | -0.96 (0.22) | 0.03 (0.27) | 7.87 | **0.001** | Controls/drug resistant: **<0.01**<br>Controls/drug responsive: > 0.90<br>Drug resistant/drug responsive: **<0.01** |
| **Psychomotor speed** |  |  |  |  |  |
| TMT A | 1.09 (0.28) | 0.12 (0.18) | 10.61 | **<0.001** | Controls/drug resistant: **<0.001**<br>Controls/drug responsive: 0.90<br>Drug resistant/drug responsive: **0.001** |
| **Verbal memory** |  |  |  |  |  |
| Verbal learning | -0.81 (0.20) | -0.43 (0.19) | 6.99 | **<0.01** | Controls/drug resistant: **<0.01**<br>Controls/drug responsive: 0.70<br>Drug resistant/drug responsive: 0.30 |
| Verbal recall | -0.03 (0.17) | -0.09 (0.15) | 0.19 | 0.8 |  |

Neuropsychological test scores are presented in mean *z*-scores (standard deviation) after standardization in relation to the respective control group (in total *n* = 59).
Abbreviations: MANCOVA, multivariate analysis of covariance; TMT, trail making test; FDR, false discovery rate
Note: bold indicates statistical significance.
\*Post-hoc *p* values are calculated from residualized metrics after covarying for age, sex and centre.

#### Neuropsychological domains

The first PCs for executive function and verbal memory had eigenvalues of 3.16/2.01 and explained 56.02%/89.62% of the total variance, respectively. Tables displaying the loadings of both PCAs are provided in the Supplemental Table 2. MANCOVA revealed a significant effect of group on cognitive performance based on the cognitive composite scores of executive function and verbal memory (Wilk’s lambda = 0.82, *F* = 6.44, *p* <0.01) covarying for age, sex and centre. ANCOVA with age, sex and centre as covariates showed significant group differences in both executive function and verbal memory (*p*_FDR_ <0.001 and *p*_FDR_ <0.05, respectively). Post-hoc t-tests adjusted calculated on residualized metrics after adjust for age, sex and centre showed worse executive function performance in drug resistant individuals against controls (*p*_Bonferroni_ <0.001) and drug responsive individuals (*p*_Bonferroni_ <0.01), whereas no difference was found between drug sensitive patients and controls. Concerning performance in verbal memory, no inter-group significant differences were found after correction for multiple comparisons. Repeated ANCOVA covarying additionally for either HADS-A or HADS-D did not alter the significance of results. The effects of group in domain-specific neuropsychological performance are summarized in Fig. 2 and provided in more detail in Table 3.

**Figure 2.**
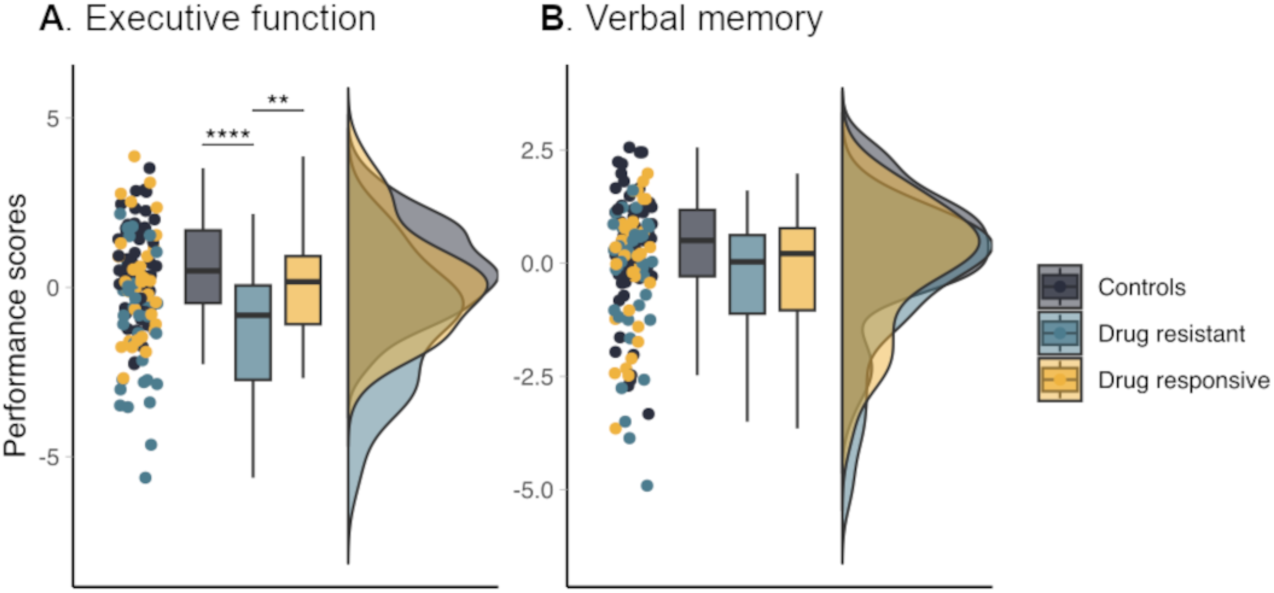
Performance in neuropsychological domains across groups. Raincloud plots show the distribution of composite cognitive construct scores representing (A) executive function and (B) verbal memory performance across healthy controls and individuals with drug resistant and drug responsive JME. In A-B, asterisks refer to *p*-values for Bonferroni-corrected, age- and sex-adjusted *post-hoc* t-tests calculated on the residuals from the analysis of covariance (ANCOVAs). Statistical details are provided in Table 3. \*\**p* <0.01, \*\*\**p* <0.001, \*\*\*\**p* <0.0001.

**Table 3.** Performance in neuropsychological domains across groups (composite scores)

|  | **Controls (*n* = 59)** | **Drug resistant (*n* = 36)** | **Drug responsive (*n* = 33)** | **F Statistic** | ***P*<sub>FDR</sub> value** | **Post hoc *p* values\***<br>**(Bonferroni corrected)** |
| --- | --- | --- | --- | --- | --- | --- |
| Executive Function | 0.61 (0.17) | -1.13 (0.35) | 0.16 (0.28) | 13.11a | **<0.001** | Controls/drug resistant: **<0.001**<br>Controls/drug responsive: 0.90<br>Drug resistant/drug responsive: **<0.01** |
| Verbal Memory | 0.33 (0.17) | -0.44 (0.27) | -0.01 (0.25) | 4.51 | **<0.05** | Controls/drug resistant: 0.30<br>Controls/drug responsive: 0.90<br>Drug resistant/drug responsive: 0.20 |

Principal component analysis derived composite cognitive mean scores (standard deviation) respective to executive function and verbal memory are presented).
Abbreviations: MANCOVA, multivariate analysis of covariance; FDR, false discovery rate.
Note: bold indicates statistical significance.
\*Post-hoc *p* values are calculated from residualized metrics after covarying for age, sex and centre.

### Role of clinical disease characteristics

Controlling for age at onset and total ASM load, epilepsy duration correlated significantly with both executive function and verbal memory (*r* = -0.31, *p*_FDR_ < 0.05; and *r* = -0.32, *p*_FDR_ < 0.05 respectively). This indicates that longer disease duration is associated with worsening performance in both neuropsychological domains independent of onset timing and ASM load. We did not find a significant correlation between both cognitive domains and age at onset while controlling for disease duration.

### Correlation between neuropsychological domains and cortical surface features

Complete neuropsychological assessment and structural brain imaging was available for 35 individuals with drug resistant JME, 31 with drug sensitive disease and 58 healthy controls.

#### Executive function

In the drug resistant group, we observed a correlation of worse performance in executive function with greater surface area within the posterior cingulum and precuneus bilaterally, the medial temporal lobe (right more than left) as well as the lateral aspects of the temporal and frontal in a diffuse symmetrical distribution (all *p*_FWE_ <0.05) after controlling for disease duration and total ASM load (see Fig. 3A). In the drug responsive and control groups, no vertices survived familywise correction for the same partial correlation analysis.

**Figure 3.**
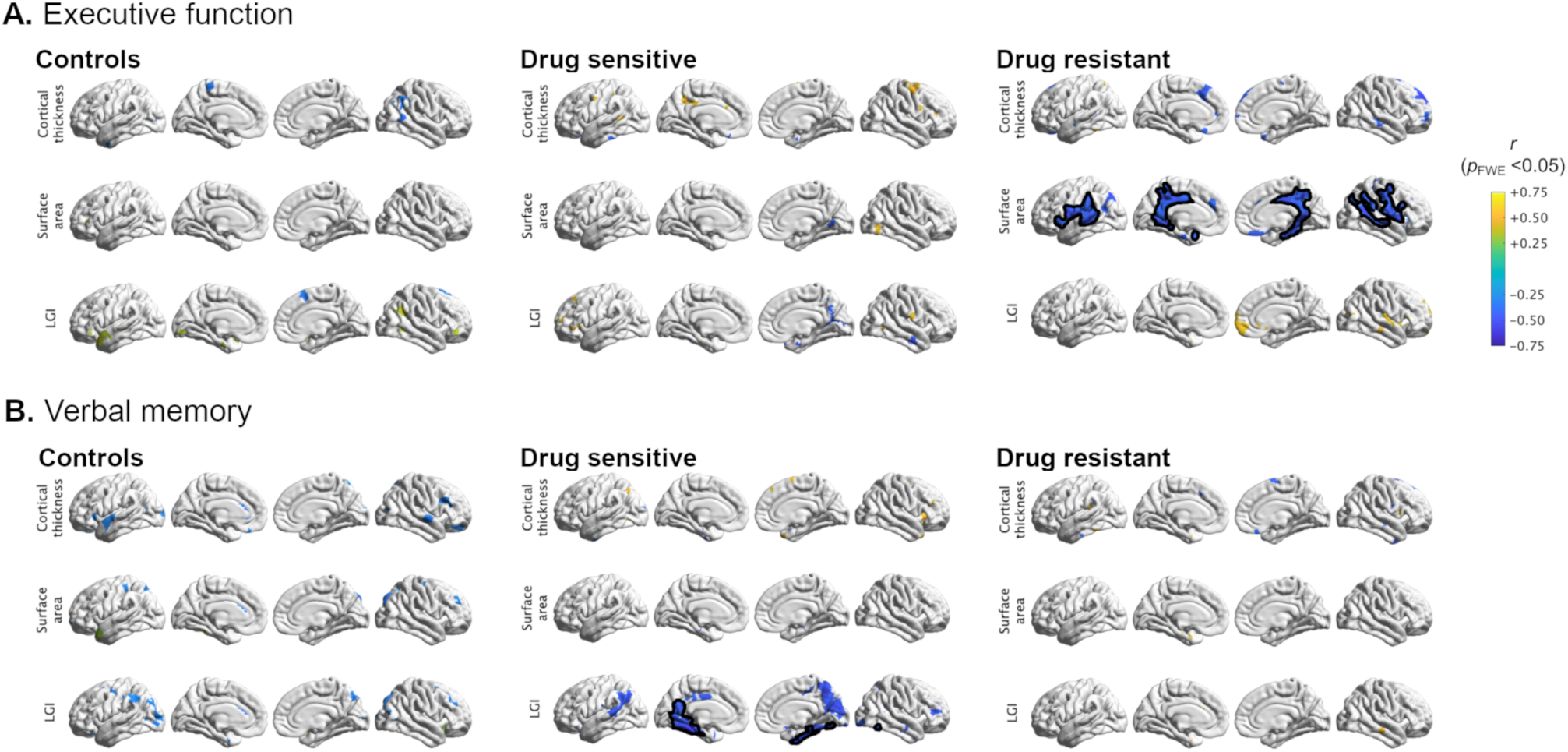
Correlation between neuropsychological domains and cortical surface features. Vertex-wise partial *Pearson* correlation plots between the composite cognitive constructs for executive function (A), verbal memory (B) and the cortical surface features (cortical thickness, surface area and local gyrification index (LGI)). Partial correlation analysis used age and sex as covariates for healthy controls, and disease duration, sex and total ASM load for the drug sensitive and drug resistant groups. Significant clusters that survived multiple comparisons corrections using random field theory at *p*_FWE_ <0.05 are outlined in black. Clusters are color-coded according to the partial Pearson correlation coefficient (*r*) (see color bar).

#### Verbal memory

When investigating the association between verbal memory performance and surface metrics, partial correlation analysis highlighted worse verbal memory with greater LGI of the medial temporal lobes in the drug sensitive group, after covarying for disease chronicity and ASM load (*p*_FWE_ <0.05) (see Fig. 3B). No significant correlations were detected in the control and drug resistant group.

## Discussion

In this cross-sectional MRI study, we observed increased white matter surface area and folding complexity with left-sided predominance in individuals with drug resistant JME compared to those with drug sensitive JME and healthy controls. Individuals with drug resistant JME also showed more pronounced on executive function tests, the extent of which correlated with the severity of identified structural changes.

Cortical folding, or gyrification, is a dynamic developmental process in which the cerebral cortex expands in volume and space, accompanied by complex tissue folding.^48–50^ Closely linked to the gyrification process is the white matter surface area, a well-established developmental marker that reflects the tangential expansion of cortical columns within a region, contributing to the extent of cortical folding.^51,52^ Abnormalities in cortical expansion and, consequently, gyrification, have repercussions on the resulting system-level connectivity^53^ and can manifest clinically in neuropsychiatric and neurological disorders. Notably, studies in children with autism spectrum disorder^39,41^ and in primates with prenatal brain injury^54^ showed an association between locally altered gyrification patterns and underlying decreased structural and functional connectivity.

Previous large-scale MRI cohort studies and meta-analyses did not identify a relationship between the identified structural abnormalities and disease severity in IGE syndromes. In our vertex-wise univariate analysis, we highlighted disease-related abnormalities in cortical surface area and folding complexity by comparing healthy controls and people with JME. In line with our previous study^19^, white matter surface area was increased in the patient group bilaterally and cortical thinning was limited to the premotor and medial prefrontal cortex on the left-side. We then investigated the structural phenotype associated with a more severe clinical course by comparing individuals with drug resistance to those with controlled disease. Clinical severity was associated with increased white matter surface area in multiple regions including the prefrontal, parietal and temporal lobes with left-sided predominance, whereas increased folding complexity as represented by LGI was limited to the left temporal lobe. As previously reported, white matter surface area and gyrification are not only genetically determined cortical features, the former is in fact also an established endophenotype in JME.^19,55^ We thus hypothesize that the abnormalities of cortical architecture observed in refractory disease reflect a spectrum of underlying post-migrational developmental anomalies which impair the network circuitry and ultimately contribute to the observed clinical phenotype with drug resistance and cognitive impairment. Our findings thus support a neurodevelopmental-network basis as the cause of drug resistance in IGE syndromes, opposing altered ASM kinetics as proposed earlier.^15,16,56^ Interestingly, when adjusting the surface area model for the effect of brain maturation (represented by patient age), significant surface area increase in drug resistant individuals was limited to the medial left temporal lobe. We speculate this reflects the complex interplay between abnormal neurodevelopmental trajectories specific to JME (as expressed by increased surface area) and normal brain maturation mechanisms. Normal aging possibly offsets the observed developmental abnormalities at some point in life, translating into the clinical benign course of the disease (in layman terms, patients “grow out of their disease”). This is supported by clinical data demonstrating that seizures become better controlled from around the forth decade of life onwards, when brain maturation is presumably completed.^57^

In contrast to sulco-gyral markers, evidence for altered cortical thickness patterns in IGEs is more conflicting. Recent large-scale studies showed a predilection of the precentral cortices for atrophy, not only in JME^19,58^ but also across all epilepsies.^24^ Here, we reproduced previous findings from large-scale studies in IGEs by showing grey matter loss in the left premotor areas. However, cortical thickness does not seem to be affected by disease severity in our cohort, thereby suggesting its role as a disease marker, but not of drug resistant disease.

Changes of network connectivity have been found in individuals with IGEs with uncontrolled seizures and are likely to drive a more severe clinical phenotype including cognitive impairment and psychiatric comorbidity.^59–61^ We explored the neurocognitive profile of our cohort by adopting a dimensionality reduction approach and focusing on neurocognitive domains, in line with recent literature.^62,63^ A more pronounced impairment of both executive function and verbal memory was associated with longer disease duration independently of ASM load or disease onset, thereby revealing a relationship between multi-domain cognitive performance and disease chronicity. Nevertheless, after adjusting for the effects of age, drug resistant disease was still associated with a higher difficulty in executive function tasks when compared to both drug responsive individuals and controls. These findings show consistency with previous studies as they suggest executive dysfunction as a core characteristic of the neuropsychological phenotype of drug resistant JME.^9,10^

Our whole-brain partial correlation analysis revealed a set of cortical areas comprising the posterior cingulum, the precuneus, the medial temporal lobe as well as peri-sylvian regions, where the extent of surface area abnormalities showed an association with worse executive function. The spatial distribution of this relationship largely overlapped with cortical regions that demonstrated increased surface area in drug resistant individuals when compared to drug responders. In light of these findings, we can infer that white matter surface area is not only increased in the medial temporal and prefrontal areas as part of a structural neurodevelopmental phenotype specific to drug resistant JME, but also that the extent of such phenotypic structural changes correlates with executive function performance, i.e., the more abnormal the structural phenotype (as represented by surface area increase), the higher the impairment in executive function. This relationship, however only seems to holds true for drug resistant disease. Interestingly, drug responsive disease showed a divergent cognitive-developmental profile, where increasing folding complexity in the medial temporal lobes bilaterally was associated with deficits in verbal memory, but not in executive function, despite preserved verbal memory performance in drug responsive disease. Indeed, not only is it evident that cortical architectural changes reflect widespread network disorder by extending beyond the prefrontal lobe to the medial temporal and parietal areas, among others, but they do so in seemingly different patterns between drug resistant and drug responsive JME. This may reflect distinct network reorganization and divergent neurodevelopmental trajectories between both disease subgroups, further emphasizing the structural heterogeneity of JME and the possibility that we are looking at different diseases. We controlled for a potential influence of ASM-related effects on cortical morphology by covarying these analysis to the total ASM load.

Our study has limitations. Given the cross-sectional design, we cannot prove whether the observed structural abnormalities are causative changes, consequences of seizure activity, prolonged drug treatment or multifactorial. Epilepsies are associated with dynamic structural changes and therefore warrant longitudinal long-term studies to capture the spatiotemporal evolution of cortical architecture. Another limitation arises from the complex interplay of anti-seizure medication and recurrent seizure activity on cognition, which adds increased difficulty in identifying a distinct neuropsychological profile in drug resistant JME. Furthermore, pseudo-drug resistance can occur in up to 20% of patients with JME and is challenging to account for.^64^ Due to lacking information regarding patient adherence to medication, some individuals may have been misclassified as drug resistant in our study.

To conclude, we identified a distinct cognitive-developmental phenotype for drug resistant JME, supporting a neurodevelopmental basis for disease severity in IGE syndromes. We provide new insights into the pathophysiology of IGEs by delivering a neurodevelopmental framework on which future genetic, imaging, and clinical studies can build on. Such studies will contribute to the development of risk stratification strategies and better phenotypical prediction models, ultimately with the goal to improve patient care through timely and tailored pharmacological and cognitive interventions.

## Data availability

The data supporting the findings of this study are available from the corresponding author upon reasonable request. Study datasets are not made publicly available due to ethical and data protection restrictions.

