## Supplementary Material for "Imaging Drug Resistance in Juvenile Myoclonic Epilepsy with MRI-derived Cortical Markers"

### Supplemental Material

**Supplemental Table 1**. Demographics in healthy controls and people with JME

**Appendix 1**. Neuropsychological tests and self-assessment questionnaires

**Supplemental Table 2**. Factor loadings for neurocognitive domain PCAs

**Supplemental Figure 1**. Effect of age and disease duration on cortical markers

**Supplemental Table 1. Demographics in healthy controls and people with JME**

| | **Controls** (*n*=78) | **People with JME** (*n*=79) | ***p* value** |
|---|---|---|---|
| Age, median [IQR], y | 26 [22, 32] | 28 [24, 35] | 0.055 |
| Female, No. (%) | 44 (56%) | 43 (54%) | 0.929 |
| From CDK, No. (%) | 59 (76%) | 51 (65%) | 0.129 |
| HADS-A, median [IQR] | 3.0 [1.8, 5.9] | 5.0 [3.0, 0.0] | **<0.001** |
| HADS-D, median [IQR] | 2.0 [1.0, 3.0] | 4.0 [1.0, 5.8] | **<0.001** |

CDK, Christian-Doppler University Hospital; HADS, Hospital Anxiety and Depression Score - Anxiety/Depression; JME, juvenile myoclonic epilepsy; IQR, interquartile range;

**Appendix 1. Neuropsychological tests and self-assessment questionnaires**

| **Neuropsychological domains** | **CDK Salzburg** | **UCL London** |
|---|---|---|
| General intelligence | Mehrfachwortschatztest Form B<br>Matrix Reasoning Test (WAIS-III) | National Adult Reading Test |
| Attention & psychomotor speed | Trail-Making-Test A&B | Stroop Color Test |
| Verbal Memory | Verbaler Lern- und Merkfähigkeitstest (German version of the Auditory Verbal Learning Test) | List learning subtest (NAB) |
| Working memory | Digit Span forward<br>Digit Span backward<br>Wisconsin Card Sorting Test | Digit Span forwards<br>Digit Span backwards<br>Stroop Colour Word Test<br>Mental Arithmetic (WAIS-III) |
| Expressive Language | | McKenna Graded Naming Test |
| Verbal Fluency | Regensburger Wortflüssigkeitstest | Controlled Oral Word Association Test |
| Verbal Comprehension | Sentence Reading Test | Vocabulary and Similarities subtests (WAIS-III) |
| Emotion Recognition | Facial Expressions of Emotion: Stimuli and Test | |
| Social cognition / Theory of Mind | Reading the Mind in The Eyes Test<br>Faux Pas Test<br>Empathy quotient[a]<br>Interpersonal Reactive Index[a] | |
| Other self-assessment questionnaires | Hospital anxiety and depression scale (German version)[a]<br>Toronto Alexithymia Scale 26[a] | Hospital anxiety and depression scale[a] |

[a]self-assessment questionnaires; Abbreviations: PMU, Paracelsus Private Medical University; UCL, University College London; NAB, Neuropsychological Assessment Battery; WAIS, Wechsler Adult Intelligence Scale;

CDK, Christian-Doppler University Hospital, Salzburg; UCL, University College London; NAB, Neuropsychological Assessment Battery; WAIS, Wechsler Adult Intelligence Scale;

**Supplemental Table 2. Factor loadings for cognitive domain PCAs**

| | PC1 | PC2 | PC3 | PC4 |
|---|---|---|---|---|
| **Executive function** | | | | |
| TMT B-A | -0.609 | 0.788 | -0.027 | -0.085 |
| Digit span | 0.234 | 0.213 | 0.949 | -0.010 |
| Phonemic fluency | 0.494 | 0.289 | -0.194 | -0.797 |
| Semantic fluency | 0.575 | 0.500 | -0.248 | 0.598 |
| **Verbal memory** | | | | |
| Verbal learning | 0.739 | 0.674 | | |
| Verbal recall | 0.674 | -0.739 | | |

**Abbreviations**: PCA, principal component analysis; PC, principal component; TMT, trail making test

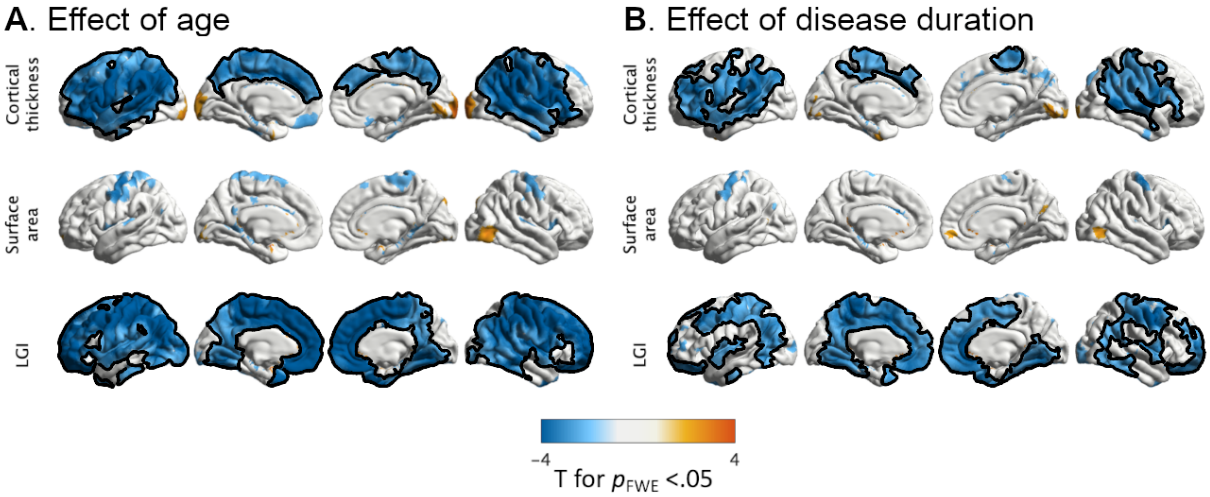


**Supplemental Figure 1. Effect of age and disease duration in cortical markers**

Univariate group analysis shows the negative effect of (A) age in the whole cohort (*n* = 157) and (B) disease duration in cortical thickness, surface area and local gyrification index (LGI) in 79 individuals with JME. The vertex-wise general linear models are two-tailed. Clusters are color-coded according to the *T* statistic (see color bar). Clusters that survived multiple comparisons correction using random field theory at $p_{FWE}$ <0.05 are outlined in black.
